# A randomized trial shows dose-frequency and genotype may determine the therapeutic efficacy of intranasal oxytocin

**DOI:** 10.1101/2020.08.23.20180489

**Authors:** Juan Kou, Yingying Zhang, Feng Zhou, Cornelia Sindermann, Christian Montag, Benjamin Becker, Keith M Kendrick

## Abstract

**Background:** The neuropeptide oxytocin is proposed as a promising therapy for social dysfunction by modulating amygdala-mediated social-emotional behavior. Although clinical trials report some benefits of chronic treatment it is unclear whether efficacy may be influenced by dose frequency or genotype.

**Methods:** In a randomized, double blind, placebo-controlled pharmaco-fMRI trial (150 male subjects) we investigated acute and different chronic (every day or on alternate days for 5 days) intranasal oxytocin (24IU) effects and oxytocin receptor genotype-mediated treatment sensitivity on amygdala responses to face emotions. We also investigated similar effects on resting state functional connectivity between the amygdala and prefrontal cortex.

**Results:** A single dose of oxytocin reduced amygdala responses to all face emotions but for threatening (fear and anger) and happy faces this effect was abolished after daily doses for 5 days but maintained by doses given every other day. The latter dose regime also enhanced associated anxious-arousal attenuation for fear faces. Oxytocin effects on reducing amygdala responses to face emotions only occurred in AA homozygotes of rs53576 and A carriers of rs2254298. The effects of oxytocin on resting state functional connectivity were not influenced by either dose-frequency or receptor genotype.

**Conclusions:** Infrequent chronic oxytocin administration may be therapeutically most efficient and its anxiolytic neural and behavioral actions are highly genotype-dependent in males.

## Introduction

Intranasal oxytocin (OT) has been proposed as novel treatment to attenuate social dysfunctions and anxiety in autism spectrum disorder, social anxiety and schizophrenia (Kendrick et al., 2017; Meyer-Lindenberg et al., 2011; Young and Barrett, 2015) as well as an augmentative strategy to facilitate fear extinction (Eckstein et al., 2015) and working memory performance (Zhao et al., 2018). Following an initial landmark study in humans demonstrating that OT reduces amygdala responses to threatening (fear and angry) face stimuli (Kirsch et al., 2005), subsequent preclinical research has consistently reported that its effects on amygdala function (attenuated fear reactivity and increased amygdala-prefrontal intrinsic connectivity - Domes et al., 2007; Eckstein et al., 2017; Kendrick et al., 2017; Meyer-Lindenberg et al., 2011; Quintana et al., 2016; Spengler et al., 2017; Sripada et al., 2013; Wang et al., 2017) may represent its primary therapeutic-relevant neural mechanism of action.

The proposed neural mechanisms of OT have been established using acute single dose administration protocols, with a dose-response study demonstrating 24 International Units (IU) as optimal for reducing amygdala responses to fear stimuli using conventional intranasal spray devices (Spengler et al., 2017). To date however, no human studies have demonstrated whether chronic treatment protocols can produce enhanced functional effects and initial clinical trials using chronic (twice daily) administration over periods of a week or longer have reported inconsistent, or at best modest therapeutic efficacy on social-emotional dysfunctions (Guastella et al., 2015; Halverson et al., 2019; Jarskog et al., 2017; Kendrick et al., 2017; Watanabe et al., 2015; Yamasue et al., 2018).

G-protein-coupled neuropeptide receptors typically exhibit internalization following even a relatively short period of constant exposure to their target peptides and cannot subsequently respond (desensitized) until they are recycled back the surface of the cell membrane (Pierce et al., 2002; Lohse and Hofmann, 2015). Growing evidence from both in vitro and animal studies suggest there may be extensive internalization and subsequent desensitization of OT receptors (OXTR) following repeated OT administration (Smith et al., 2006; Stoop, 2012). In terms of neural OXTR receptors those in the amygdala may be particularly susceptible to desensitization (Terenzi and Ingram, 2005) and this region critically mediates many of the effects of intranasal OT on social cognition and anxiety (Kendrick et al., 2017). Moreover, rodent models have reported that in contrast to single doses, chronic administration of OT can actually produce social impairment (Bales et al., 2013; Du et al., 2017; Huang et al., 2014) in the context of reduced receptor expression in the amygdala and nucleus accumbens (Du et al., 2017). Furthermore, an initial study reported that chronic versus single administration of OT results in divergent neurochemical changes in both the rodent and human frontal cortex (Benner et al., 2018), emphasizing high translational relevance of the preclinical animal models for clinical trials employing chronic treatment protocols. Thus, empirically evaluated optimal treatment protocols for chronic intranasal OT administration in humans are urgently needed to determine its therapeutic potential.

A further unresolved issue is that different OT receptor (OXTR) genotypes have been associated with social behaviors and may influence sensitivity of behavioral and neural responses to intranasal OT. In particular, OXTR polymorphisms rs53576 and rs2254298 have been associated with autism (Cataldo et al., 2018), deficits in social and emotional processing and anxiety (Jurek and Neumann, 2018; Parker et al., 2014; Yang et al., 2017) and individual variations in behavioral and neural responses to intranasal OT (Chen et al., 2015; Feng et al., 2015). Indeed, variants of rs2254298 may represent a trans-diagnostic biomarker for social dysfunctions (Brüne, 2012). It is therefore imperative to establish whether OXTR polymorphisms could influence sensitivity to effects of intranasal OT.

While single doses of OT can produce numerous functional effects, two of key importance are attenuated amygdala responses to face emotions, particularly fear, and increased resting state functional connectivity between the amygdala and prefrontal cortex (Domes et al., 2007; Eckstein et al., 2017; Kirsch et al., 2005; Koch et al., 2016; Quintana et al., 2016; Spengler et al., 2017; Sripada et al., 2013). While most task studies have focused on the effects of OT on amygdala responses to fear faces using a variety of different task paradigms some studies have reported valence independent effects of OT (Domes et al., 2007; Koch et al., 2016; Quintana et al., 2016). Both these task and resting state neural markers have been primarily associated with attenuated anxiety in terms of reduced responses to social threat and enhanced top-down control of emotion (Zhao et al., 2019), although effects of OT on the amygdala may influence a range of social cognition domains (Kendrick et al., 2017; Meyer-Lindenberg et al., 2011; Young and Barrett, 2015).

To help determine optimal treatment protocols for chronic intranasal OT administration the current pre-registered double-blind, randomized between subject placebo (PLC)-controlled pharmacological neuroimaging trial therefore aimed to replicate previous findings for effects of a single dose of OT (24IU) on reduced amygdala responses to fear or other face emotions (Domes et al., 2007; Kirsch et al., 2005; Koch et al., 2016; Quintana et al., 2016; Spengler et al., 2017; Wang et al., 2017) in a large sample at the whole brain level and on increasing resting-state functional connectivity between the amygdala and prefrontal cortex (Eckstein et al., 2017; Sripada et al., 2013). Next, the primary outcome of the trial was to establish whether different repeat dosing protocols (daily, OT_5_ group, or every other day, OT_3_ group, for 5 days) modulate amygdala-centered neural and behavioral responses and resting state functional connectivity. As a secondary outcome, we further aimed to establish if effects of OT on primary outcome measures were influenced by OXTR genotype, focusing on the rs53576 and rs2254298 polymorphisms most frequently associated with social function and individual variations in intranasal OT sensitivity.

## Material and Methods

The main study objectives were to investigate both the effects of acute (single dose) and repeated doses (daily or every other day for 5 days) of 24IU OT versus PLC on two primary outcomes: (1) amygdala and behavioral responses to face emotions (particularly fear faces) and (2) resting state functional connectivity between the amygdala and frontal cortex. We investigated modulatory influences of OXTR rs53576 and rs2254298 polymorphisms on the sensitivity to intranasal OT as a secondary outcome.

### Participants

A total of 150, right-handed healthy adult male subjects were enrolled according to common inclusion and exclusion criteria for human OT-administration studies (see **SI**). Given that methods for estimating power for whole brain task-based fMRI analyses are limited (Poldrack et al., 2017), subject number was initially chosen on the basis of recent large-scale studies investigating dose effects of OT (Spengler et al., 2017 n = 111 males; Lieberz et al., 2019, 90 females). A priori power estimates for behavioral analyses were calculated using Webpower (Zhang and Yuan, 2018) giving a range of 60-80% power for ANOVA main and interaction effects and post-hoc tests. For full details see SI. Calculating power for fMRI experiments is more problematic and likely to be much lower, although since the amygdala was our only pre-registered ROI of interest, and the majority of fMRI post-hoc analyses used a single amygdala ROI, this avoids the problem of significantly reduced power when undirected whole brain analyses are performed (see Cremers et al., 2017). The current chosen sample size is also comparable to the previous largest fMRI studies investigating task-related effects of OT (Spengler et al., 2017; Lieberz et al., 2019). Three subjects failed to meet inclusion criteria (were currently taking medications) and 9 subjects were excluded due to failure to complete the study or excessive head movement (see **Fig. S1**). Subjects were randomly (computer-generated randomization) assigned to repeated intranasal treatment (single daily dose on five consecutive days) of (1) placebo (PLC; n = 46, M ± SD, 22.46± 2.3 years), (2) oxytocin (OT_5_; n = 49, M ± SD, 21.78 ± 2.3 years), or interleaved OT and PLC (OT on days 1, 3, 5, PLC on days 2, 4; OT_3_; n = 43, M ± SD, 21.02 ± 2.0 years) (see **SI**). To ensure compliance all subjects were supervised during self-administration of all nasal sprays. Groups did not differ on a number of potential confounders (see **SI** and **Table S4**). The study was approved by the local ethics committee (Institutional Review Board, University of Electronic Science and Technology of China) and subjects provided written informed consent. The study was in accordance with the latest revision of the Declaration of Helsinki, pre-registered at Clinical Trials.gov (NCT03610919 - https://clinicaltrials.gov/ct2/show/NCT03610919) and in line with recommendations for trials in psychological experiments (Guidi et al., 2018) (see **Fig. S1** for Consort flow diagram). Subjects received 500 RMB monetary compensation for participation.

### Experimental procedures

The study employed a double-blind, randomized, placebo-controlled, between-subject design. The OT and PLC sprays used in the 3 groups were supplied by Sichuan Meike Pharmaceutical Co. Ltd, Sichuan, China in identical dispenser bottles containing identical ingredients (glycerine and sodium chloride) other than OT. In line with recommended guidelines experiments started 45 minutes after intranasal administration (Guastella et al., 2013). Recent studies have confirmed that intranasal OT enters the brain (Lee et al 2018), increases cerebrospinal fluid and blood concentrations (Striepens et al., 2013) and produces activity changes in brain regions expressing OXTRs in humans at around 45 minutes after administration (Paloyelis et al., 2016; Quintana et al 2019). In post-treatment interviews on days 1 and 5 subjects could not identify better than chance whether they had received OT or PLC (χ^2^ < 0.1, ps > 0.2, **Table S4**). For OXTR genotyping subjects provided buccal swaps on the 1^st^ day for analysis of OXTR rs2254298, rs53576 SNPs (see **SI** and (Montag et al., 2017)).

For the implicit face-emotion processing task 208 grayscale facial stimuli displaying happy, neutral, angry or fearful facial expressions (n = 26 per category, 50% female) were used (two different stimulus sets - see **SI** for details).

### Primary outcomes and analysis plan

Face emotion-related amygdala responses were assessed using the event-related implicit face processing fMRI paradigm on treatment days 1 and 5. Intrinsic amygdala connectivity was assessed by means of resting state fMRI assessments before the task-paradigm. Valence, arousal and intensity ratings (scale: 1-9) for the facial stimuli were collected immediately after MRI acquisition as additional behavioral outcomes (further details see **SI**). In line with the main aim of the study changes in amygdala face emotion reactivity and amygdala intrinsic connectivity between the 1^st^ and 5^th^ day served as primary outcome measures. Changes in valence, arousal and intensity ratings for the different face emotions represented an associated behavioral outcome measure.

### Imaging acquisition and analysis

Blood oxygenation level-dependent contrast functional images were acquired using standard sequences on a 3T GE MR750 system. High resolution T1-weighted structural MRI data were acquired on both 1^st^ and 5^th^ days to improve normalization. MRI data was preprocessed using validated procedures in SPM12 (Friston et al., 1994) (Statistical Parametric Mapping; http://www.fil.ion.ucl.ac.uk/spm) and Data Processing Assistant or Resting-State fMRI (Yan, 2010) (DPARSFA; http://rfmri.org/DPARSF) (for details see **SI**). First level General Linear Models (GLM) for the task-related fMRI data included separate regressors for the four emotional conditions, gender identity rating period and 6 movement parameters and appropriate contrasts were subjected to a second level random effects analysis.

### Primary outcomes: The effects of OT on amygdala and behavioral face emotion reactivity and intrinsic connectivity

Differences in amygdala and behavioral responses to face emotions were examined using mixed ANOVAs with treatment group (PLC, OT_3_, OT_5_) as between-subject and time point (1^st^ and 5^th^ days) and face emotion (angry, fear, happy and neutral) as within-subject factors. Dependent variables were extracted target region amygdala responses using the template of left and right amygdala masks from the Brainnetome atlas (Fan et al., 2016) to faces (parameter estimates extracted using Marsbar, http://marsbar.sourceforge.net) and valence, intensity and arousal ratings. Post-hoc comparisons explored significant interactions for amygdala and behavioral ratings. Associations between amygdala responses and behavioral ratings were assessed using Pearson correlation.

Differences in resting state amygdala functional connectivity were determined using a voxel-wise seed-to-whole-brain ANOVA as implemented in the SPM flexible factorial design with treatment (PLC, OT_3_, OT_5_) as between-subject and time point (1^st^ and 5^th^ days) as within-subject factors and amygdala connectivity maps as dependent variable. (See data analysis flow chart in **SI**)

### Secondary outcome measures: influence of OXTR genotype

For rs2254298 subjects were divided into A carriers (AA and AG) and A non-carriers (GG) and rs53576 into G carriers (GG and GA) and G non-carriers (AA). To explore effects of genotype as a secondary outcome measure, OXTR group was included as an additional between-subject factor in the corresponding ANOVAs and amygdala reactivity to face emotions, amygdala-prefrontal cortex intrinsic functional connectivity and behavior rating response served as dependent variables.

### Control for treatment effects on brain structure

To control for potential confounding effects of single- and repeated OT-administration on brain structure, a voxel-based morphometry (VBM) analysis was conducted on the T1-weighted images acquired on both testing days using SPM12 standard procedures (Ashburner, 2007; Ashburner and Friston, 2005)(see **SI**).

### Thresholding and statistical implementation

In line with our regional hypothesis the analysis of the implicit emotional face task focused on the amygdala. To account for the complex design of the trial we employed two analytic approaches: (1) A region of interest analysis (ROI) on the extracted beta estimates from atlas-based masks for the left and right amygdala (Brainnetome Atlas, Fan et al., 2016) respectively. To this end the extracted estimates were subjected to three-way ANOVAs (treatment ×time point ×face emotion) examining effects on the left and right amygdala separately. The p-value for the analysis was adopted to p<0.025 (Bonferroni corrected multiple comparison 0.05/2). (2) To facilitate a more specific localization of the effects and to determine a more specific seed mask for the seed-to-whole brain resting state functional connectivity analysis a voxel-wise small volume corrected (SVC) analysis for the entire bilateral amygdala was additionally employed. To this end a peak-level Family-wise error (FWE) small volume correction was applied to a single mask encompassing the bilateral amygdala as provided in the Brainnetome Atlas (Fan et al., 2016) (total volume of the mask, 4572.27mm^3^) (FWE p < .05). For the seed-to-whole brain resting state data analysis a whole-brain cluster-based FWE-correction with an initial cluster forming threshold of p < .001 and FWE p < .05 was employed. Behavioral and neural (extracted parameter estimates) indices were further analyzed using SPSS 22.0 with appropriate ANOVA models and Bonferroni corrected post-hoc tests. P < 0.05 two-tailed was considered significant. Partial eta squared (*η*^2^_p_) and Cohen’s *d* were computed for behavioral results as measures of effect size and SPSS outlier analysis was used to confirm the absence of statistical outliers. Full details of the post-hoc and correlation analyses are provided in the SI (Tables S7 - S11).

## Results

### Primary outcome measure: ROI analysis of acute and repeated doses on OT-evoked changes in amygdala responses to face emotions

Examination of the extracted parameter estimates from the amygdala (a priori ROI approach in accordance with trial pre-registration) revealed a significant three-way ANOVA for the right amygdala (F_(6,405)_=2.563; p<0.019), but not the left amygdala (F_(6,405)_=1.860; p=0.087). Post-hoc analyses on the extracted beta values from the right amygdala mask on the 1^st^ day revealed that both OT groups compared to PLC exhibited a suppression of right amygdala reactivity (fear: ps < 0.015, angry: ps < 0.016; happy: ps < 0.041: neutral: ps < 0.017). Analysis on the 5^th^ day revealed that the OT_3_ group exhibited a suppression of right amygdala reactivity to fear and angry faces relative to both the PLC (fear: p = 0.003, angry: p = 0.008) and OT_5_ groups (fear: p = 0.021; angry: p = 0.082) but the OT_5_ group did not (ps > 0.315, relative to PLC group). The OT_3_ group also exhibited reduced right amygdala reactivity to happy faces on the 5^th^ day compared with the PLC group (p = 0.028) but there was no significant difference between OT_3_ and OT_5_ groups (p = 0.272). There were no significant effects of OT on the 5^th^ day for neutral faces (ps>0.203, although OT_3_ vs PLC was marginal, p = 0.052). While amygdala reactivity to fear faces did not differ in the PLC group between days 1 and 5 (p = 0.356), its reduction in the OT_3_ group on day 5 was equivalent to day 1 (p = 0.538) whereas in the OT_5_ group the reduction on day 5 was significantly less than on day 1 (p = 0.035). Amygdala reactivity to angry faces decreased significantly more in the OT_3_ group on day 5 relative to day 1 (p = 0.006), although it also did so in the PLC group for both angry (p = 0.007) and neutral (p = 0.028) (**Fig. 1b-e**).

**Fig. 1.**
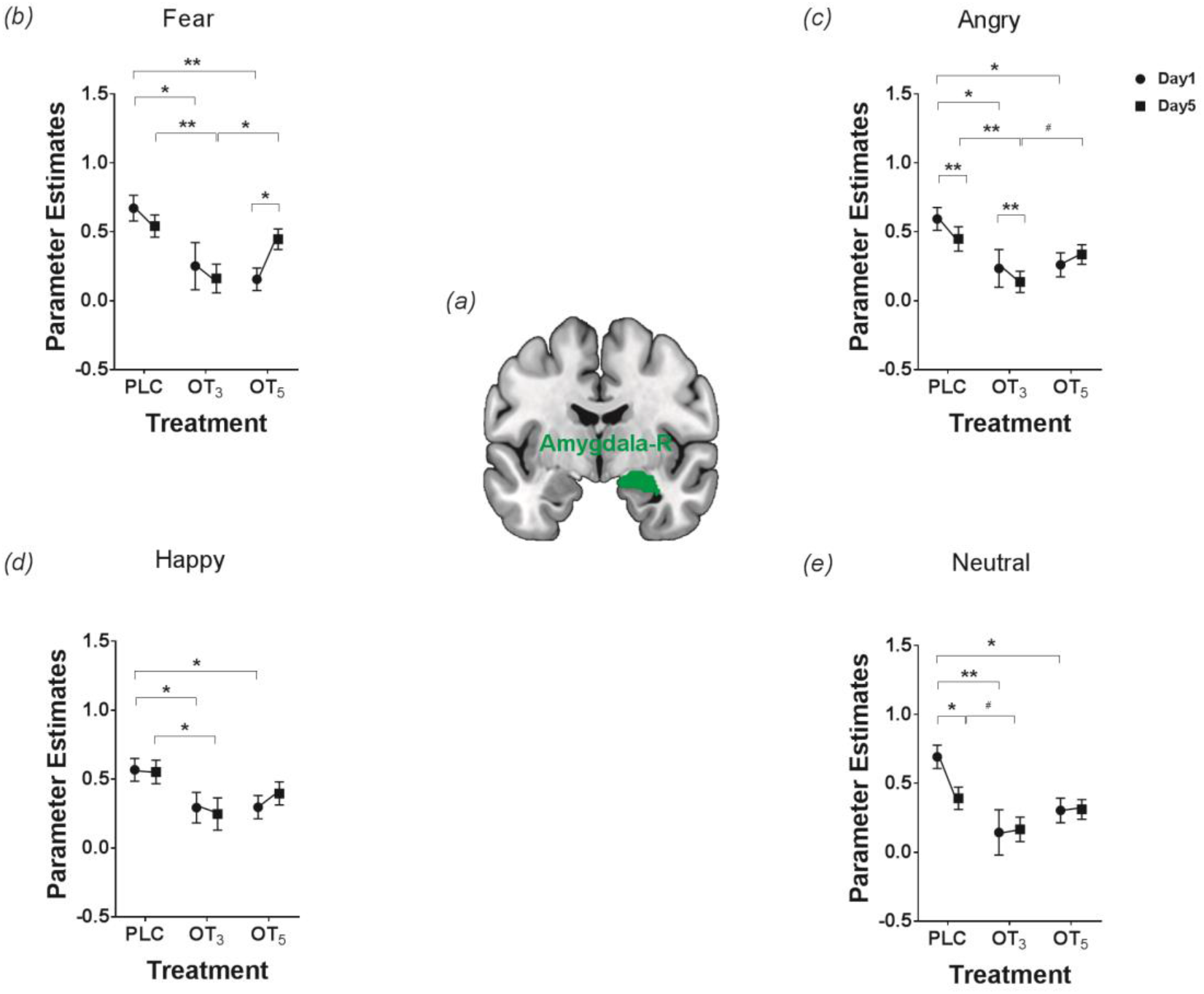
The effect of intranasal oxytocin (OT) treatment on neural responses to faces. (a) The mask of right amygdala from the Brainnetome atlas (Fan et al., 2016)(b-e) Post hoc analysis on extracted parameter estimates from the right amygdala mask for all face emotion categories on the 1^st^ and 5^th^ days.# p < 0.1, * p < 0.05, ** p < 0.01, two-tailed. Bars depict standard errors.

### Primary outcome measure: behavioral rating analysis of acute and repeated doses to face emotions

Mixed ANOVAs on the behavioral data revealed a significant main effect of treatment group on arousal ratings (arousal: F_2, 135_ = 5.06, p = 0.008, *η*^2^_p_ = 0.07) due to ratings generally being lower in the OT_3_ group. There was also a marginal significant treatment group x time point x face emotion interaction effect for arousal (F_6,405_ = 1.83, p = 0.093, *η*^2^_p_ = 0.03) and a significant one for intensity (F_6,405_ = 3.327, p = 0.004, *η*^2^_p_ = 0.05). Post-hoc analysis showed that intensity ratings were significantly reduced for fear faces in the OT_3_ group on the 5^th^ day (fear: p = 0.004 versus PLC, *d* = 0.62, 95% CI, −1.299 to −0.247, p = 0.088 versus OT_5_, *d* = 0.33, 95% CI, −0.968 to 0.068). Exploratory post-hoc analysis of arousal ratings revealed a similar reduction to fear faces on day 5 (p = 0.001 versus PLC, *d* = 0.77, 95% CI, −1.544 to −0.420, p = 0.033 versus OT_5_, *d* = 0.42, 95% CI, −1.158 to −0.051). Arousal ratings to fear faces were also decreased in the OT_3_ group on the 1^st^ day compared to the PLC group but not the OT_5_ group (arousal: p = 0.015 versus PLC, *d* = 0.60, 95% CI, 0.129 to 1.175, p = 0.355 versus OT_5_, 95% CI, −0.268 to 0.744). Arousal and intensity ratings to angry faces were reduced on day 5 compared with day 1 in the OT_3_ (arousal: p = 0.001, *d* = 0.30, 95% CI, 0.164 to 0.619, intensity: p = 0.001, *d* = 0.32, 95% CI, 0.173 to 0.641) and OT_5_ groups (arousal: p = 0.023, *d* = 0.19, 95% CI, 0.032 to 0.461, intensity: p = 0.006, *d* = 0.26, 95% CI, 0.088 to 0.526). There were no significant differences for angry, neutral and happy faces between OT and PLC groups on the 1^st^ day (ps >0.194, although OT_5_ vs PLC was marginal, p=0.051 for neutral faces) and 5^th^ day (ps >0.671, although OT_3_ vs PLC p=0.053 was marginal for angry faces). There was however a significant group difference between OT_3_ and OT_5_ on the 1^st^ and 5^th^ day for happy and neutral faces (ps<0.007) (**Fig.2**). While there was a significant 3-way interaction for valence ratings (F_6, 405_ = 2.63, p = 0.016, *η*^2^_p_ = 0.04), post-hoc analysis revealed no significant differences between OT and PLC groups on the 1^st^ (ps > 0.200) or 5^th^ days (ps> 0.173).

**Fig. 2.**
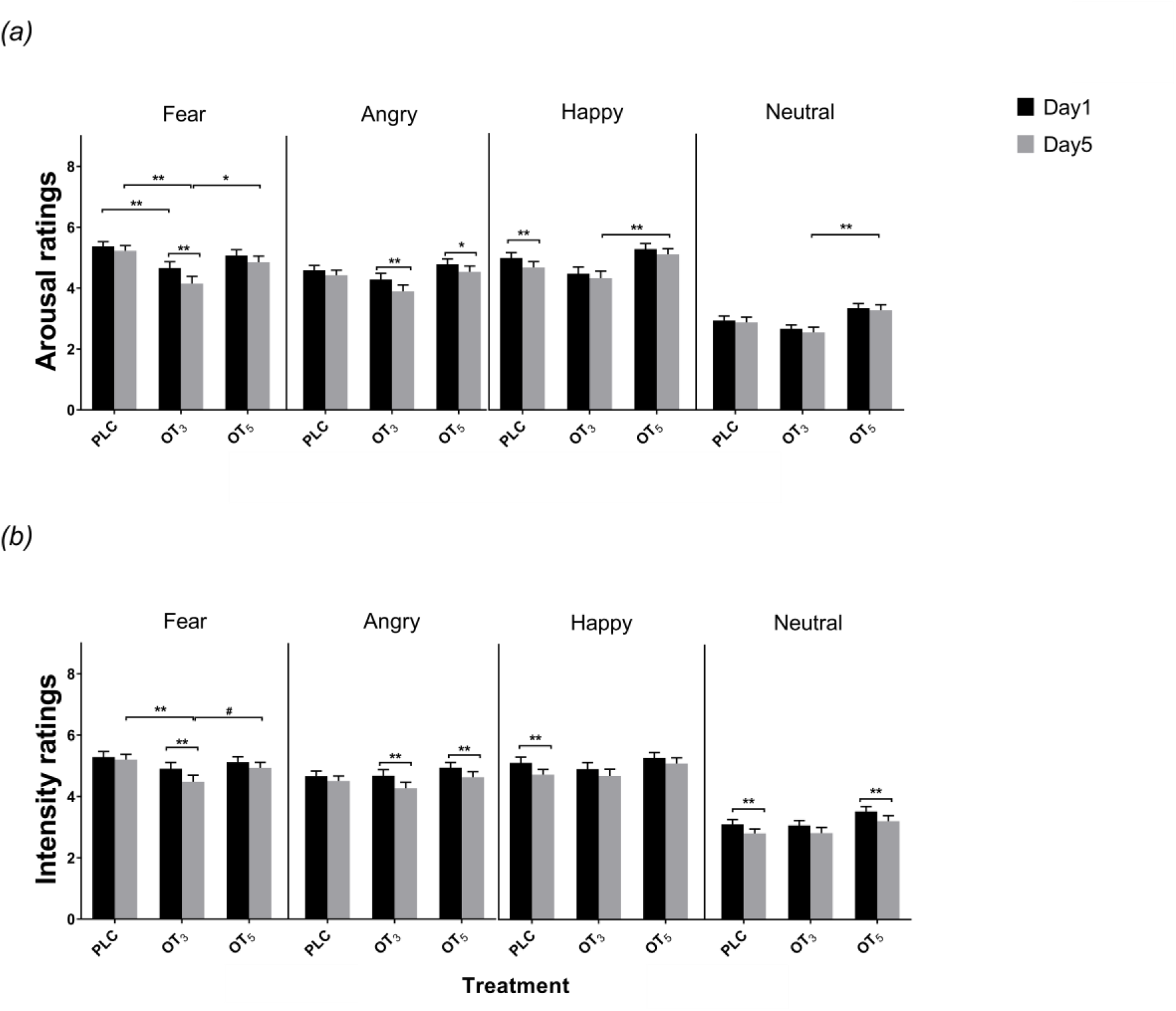
The effects of oxytocin (OT) on arousal and intensity ratings for face emotions. (a) OT decreased arousal and intensity ratings of fear faces on both the 1^st^ and the 5 days of treatment in the group receiving OT on alternate days (OT_3_) compared with placebo (PLC) and ratings were significantly reduced on day 5 relative to day 1. The group receiving OT every day (OT_5_) did not show significant effects on either day 1 or 5. No effects of OT on ratings for (b) anger, (c) happy and (d) happy face emotions. # p < 0.1, * p < 0.05, ** p < 0.01 two-tailed, Bars depict standard errors.

The whole brain voxel based morphometry analysis revealed no significant effects of acute or repeated doses of OT on gray matter volumes.

### Associations between amygdala responses and behavioral ratings to emotional faces

Associations between right amygdala activation (extracted parameter estimates from the right amygdala mask as provided by the Brainnetome atlas) and emotional arousal and intensity scores to fear faces were observed in the PLC group on the 1^st^ day (arousal: r = 0.295, p = 0.023; intensity: r = 0.455, p = 0.001) demonstrating that greater amygdala activation in response to fear faces was associated with increased anxiety. This association was absent in both OT groups on the 1^st^ day (arousal: OT_3_, r = 0.041, p = 0.398; OT_5_, r = 0.050, p = 0.366; intensity: OT_3_, r = 0.098, p = 0.265; OT_5_, r = 0.164, p = 0.130). A similar trend occurred on the 5^th^ day (arousal: PLC, r = 0.306, p = 0.019; OT_3_, r = −0.011, p = 0.471; OT_5_ group, r = −0.120, p = 0.206; intensity: PLC, r = 0.212, p = 0.079; OT_3_, r = −0.108, p = 0.246; OT_5_ group, r =-0.043, p = 0.385) (**Fig.3**). There was a similar relationship with responses to angry faces for intensity but not arousal ratings in the PLC group on the 1^st^ day. (arousal: r = 0.107, p = 0.240; intensity: r = 0.297, p = 0.023). No associations were found for responses to neutral or happy faces.

**Fig. 3.**
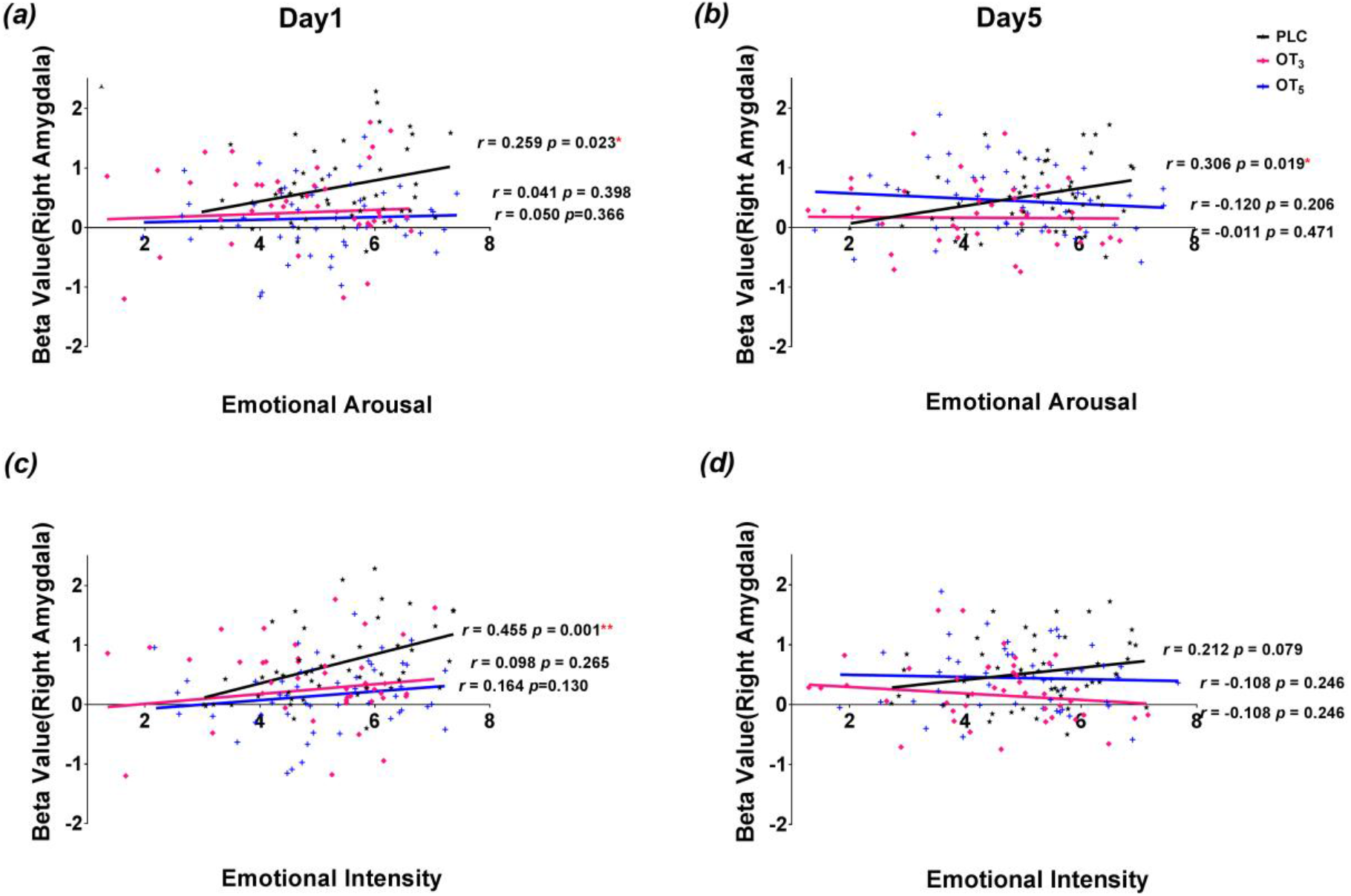
Associations between intensity and arousal ratings of fear faces and amygdala activation in the placebo (PLC) and two OT treatment groups (OT_3_ and OT) on days 1 and 5. The magnitude of right amygdala (from the Brainnetome atlas) responses was positively correlated with arousal and intensity ratings in the PLC group on day 1 but not in the OT groups. There was a similar trend on day 5. *p< 0.05. **p<0.01

### Primary outcome measure: effects of acute and repeated oxytocin doses on resting-state functional connectivity

To facilitate a more precise localization of the coordinates of amygdala for the resting state analysis a voxel-wise mixed ANOVA with treatment group (PLC, OT_3_, OT_5_) as between-subject, time point (1^st^ and 5^th^ days) and face emotion (happy, neutral, angry and fear) as within-subject factor was computed on the task-based data. This analysis revealed a significant three-way interaction effect in the right amygdala after small volume correction for the bilateral mask encompassing the entire amygdala (k = 7, p_FWE_ = 0.021, F_6,405_=4.47, peak MNI co-ordinate: x=18, y=-7, z=-16) (**Fig.4a**) confirming the findings from the ROI analysis which revealed a significant interaction effect for the right amygdala only. An amygdala seed-to-whole brain fMRI resting-state analysis employing this region as seed (6 radius sphere centered at x=18, y=-7, z=-16) by mixed-effect ANOVA revealed a main effect of treatment (OT_3_, OT_5_ and PLC) on amygdala functional coupling with the anterior prefrontal cortex (peak MNI coordinate x = −3, y =53, z = −19, F_2, 135_ = 16.55, p_FWE_ = 0.001, k = 88) (see Fig. 4a) but no treatment group x time point interaction. Right amygdala intrinsic connectivity with the prefrontal cortex was stronger in both the OT_3_ (k = 74, p_FWE_ = 0.014, x = −3, y = 53, z = −19) and OT_5_ groups (k = 127, p_FWE_ = 0.001, x = −3, y = 53, z = −19) relative to PLC. The OT_3_ and OT_5_ groups did not differ significantly on the 1^st^ (p = 0.147) or 5^th^ (p = 0.107) days (see Fig. 4b). Correlation analysis revealed no significant association between resting state and amygdala task-related changes (r=-0.08, p=0.59 in PLC group) or behavioral ratings of emotional faces (−0.085<rs<0.055, ps > 0.57).

**Fig. 4.**
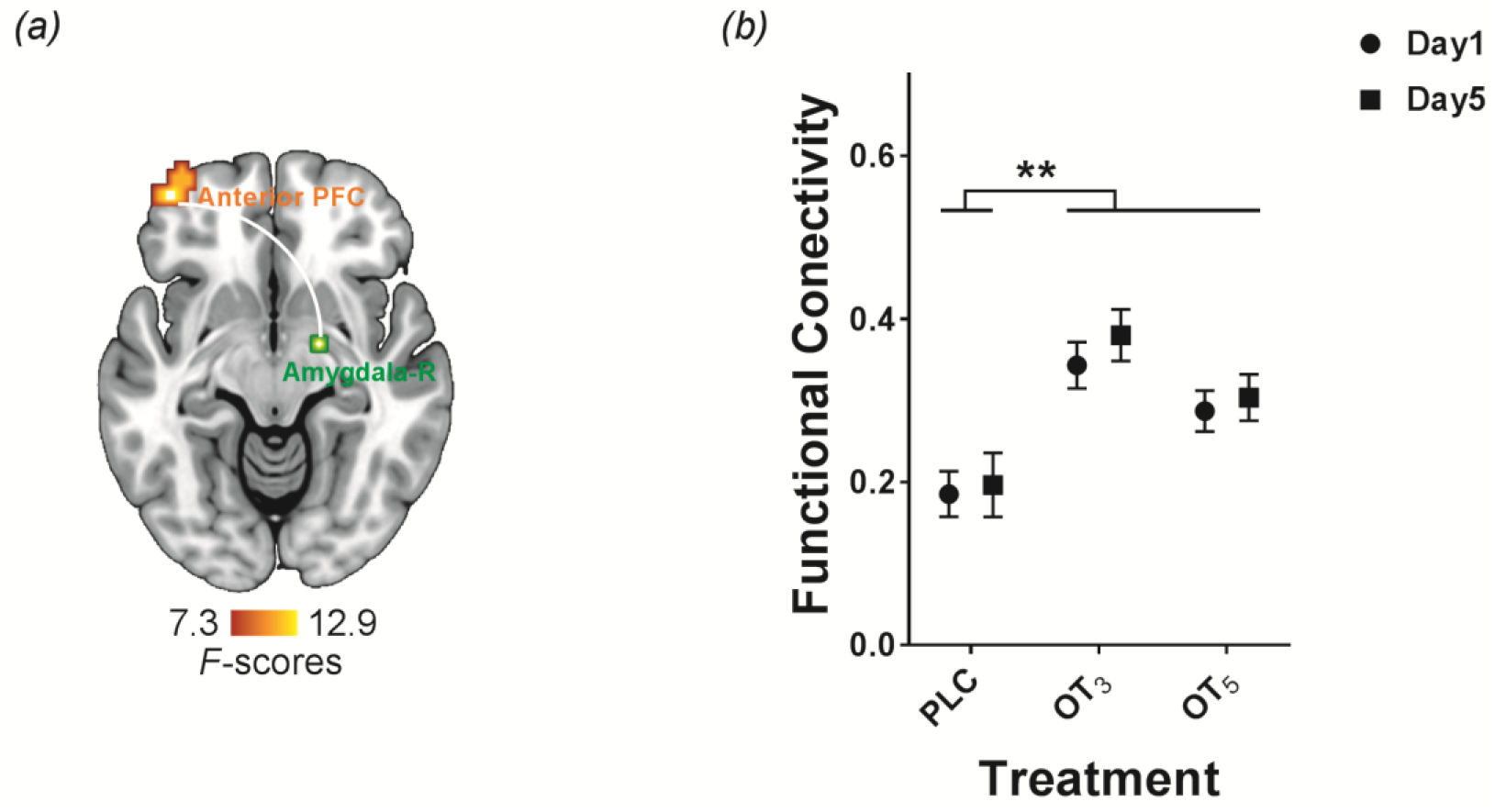
The effects of oxytocin (OT) on resting state functional connectivity (a) schematic visualization of the identified functional pathway between the right amygdala and anterior prefrontal cortex (replicating Eckstein et al., 2017). (b) Seed to whole-brain analysis of functional connectivity (FC, seed region right amygdala, 6mm sphere, x=18, y=-7, z=-16) revealed significantly increased functional connectivity between right amygdala and anterior prefrontal cortex (p_FWE_ =0.039, x = −36, y =59, z = −7) in both OT treatment groups (OT_3_ and OT_5_) on both, days 1 and 5 relative to placebo (PLC). Line graphs depict the extracted parameter estimates from right amygdala connectivity with anterior prefrontal cortex (M ± SE). * p < 0.05, ** p < 0.01 two-tailed. Bars indicate standard errors.

### Secondary outcome measure: Associations between OT-effects and OXTR genotype

The number of G-carriers and G-non-carriers of rs53576 and A-carriers and A-non-carriers of rs2254298 did not differ between the three groups and both SNPs satisfied Hardy Weinberg Equilibrium (see **Tables S2 and S3**). A four-way mixed ANOVA with treatment group and genotype as between- and time point and face emotion as within-subject factors revealed significant treatment x genotype interactions for both rs2254298 and rs53576 (rs 2254298: F_2, 105_ = 3.17, p = 0.046, rs53576: F_2, 105_ = 3.23, p = 0.044) for amygdala responses to emotional faces, although they would not survive a Bonferroni correction for 4 genotypes (i.e. p<0.0125). Post-hoc analysis showed that reduced amygdala reactivity to emotional faces in both OT groups occurred in A-carriers of rs2254298 (ps < 0.009) but not A-non-carriers (ps>0.587) and G-non-carriers of rs 53576 (ps < 0.002) but not G-carriers (ps>0.196) (see Fig. 5a-b). For behavioral ratings there were significant treatment group x genotype interactions for both arousal and intensity ratings for rs2254298 (arousal: F_2, 105_= 6.60, p = 0.002, *η*^2^_p_ = 0.11; intensity: F_2, 105_= 3.85, p = 0.024, *η*^2^_p_ = 0.07). Post-hoc analysis showed that decreased ratings for faces in the OT_3_ group only occurred in A-carriers (arousal: versus PLC: p = 0.002, *d* = 4.68, 95% CI, 0.363 to 1.524 and OT_5_ p < 0.001, *d* = 6.56, 95% CI, 0.743 to 1.904; intensity: PLC: p = 0.021, *d* = 3.40, 95% CI, 0.109 to 1.301; OT_5_: p = 0.003, *d* = 4.40, 95% CI, 0.314 to 1.507). No significant main or interaction effects involving treatment and genotype were found for resting state functional connectivity between amygdala and prefrontal cortex (rs 2254298 all ps > 0.123; rs53576 all ps > 0.566).

**Fig. 5.**
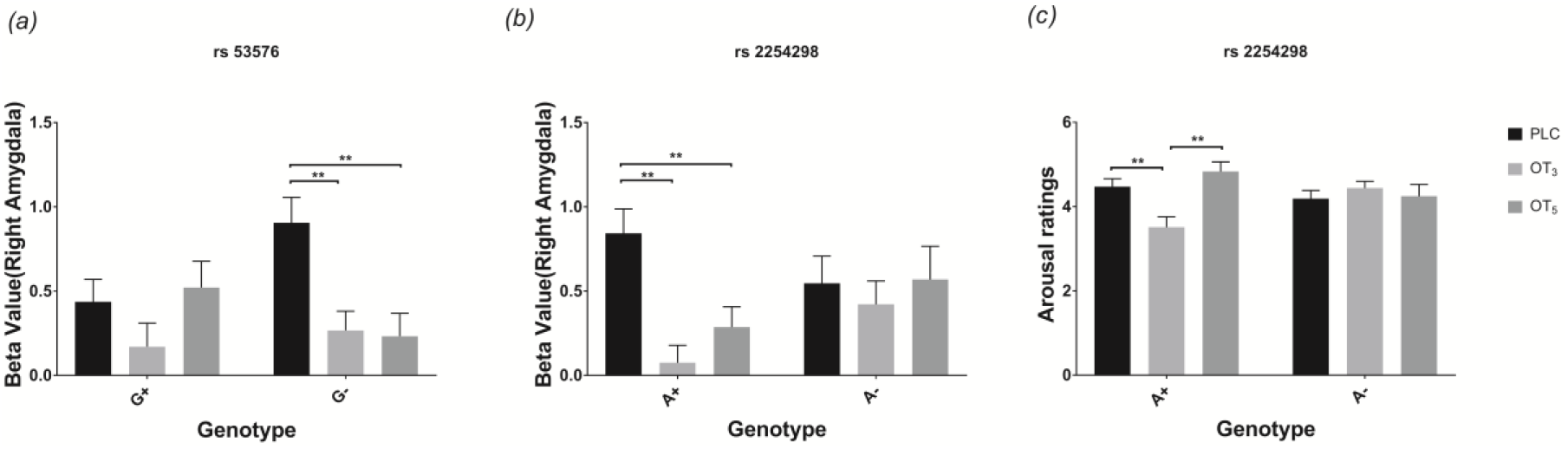
Influence of oxytocin (OT) receptor genotype on the effects of intranasal OT on right amygdala and behavioral responses to face emotions (n = 111 subjects). (a) for rs 53576 only G-non carriers (i.e. AA) showed reduced amygdala responses to faces (PLC, n_G+_=15, n_G-_=21; OT_3_, n_G+_=17, n_G-_=24; OT_5_, n_G+_=20, n_G-_=14); (b) for rs2254298 only A-carriers (i.e. AA and AG) in the groups with intranasal oxytocin (OT) treatment showed reduced amygdala responses to faces (PLC, n_A+_=20, n_A-_=16; OT_3_, n_A+_=23, n_A-_=18; OT_5_, n_A+_=20, n_A-_=14); (c) for rs2254298 only A-carriers (i.e. AA and AG) in the groups with intranasal oxytocin (OT) treatment showed reduced emotional arousal ratings ** p < 0.01, two-tailed t-test (Bonferroni corrected significance threshold of p = 0.0125). Bars indicate standard errors.

## Discussion

Overall, our findings validate OT-reductions in amygdala responses to face emotions and associated emotional intensity and arousal ratings for fear faces, together with increased resting state functional connectivity between the amygdala and prefrontal cortex as robust markers for its putative therapeutic mechanisms of action. While, these task-dependent and resting-state effects of OT show a different sensitivity to repeated intranasal doses and OXTR genotype our findings indicate that to achieve maximal effects of chronic treatment on neural and behavioral responses to face emotion stimuli and resting-state changes the optimal protocol may be to administer 24IU doses every other day rather than daily.

Our findings are highly consistent with preclinical animal models demonstrating OXTR desensitization following repeated doses of OT in some brain regions (Terenzi and Ingram, 2005; Smith et al., 2005; Stoop, 2012) and that chronic administration can reduce brain OXTR expression (Du et al., 2017) and alter patterns of neurochemical release (Benner et al., 2018). Importantly, chronic doses of OT in rodents fail to produce anxiolytic effects normally seen with single doses (Du et al., 2017) which mirrors our present observations. The apparent long-lasting desensitization effects of daily OT administration on amygdala responses to threatening and happy faces may be contributed to by dose magnitude and so possibly lower daily doses might be less problematic. We also observed some evidence for reduced amygdala and behavioral responses to repeated emotional face presentations in the PLC group in line with previous observations (Fischer et al., 2003; Wright et al., 2001), even though different sets of face stimuli were used at the two time-points. This may have served to reduce time-dependent effects of OT to some extent.

The amygdala is one of the main neural substrates mediating OT’s functional effects and its attenuation of reactivity to threatening stimuli is considered as a putative primary therapeutic mechanism of action (see Jurek and Neumann, 2018; Kendrick et al., 2017; Neumann and Slattery, 2016). However, in the current study we demonstrated similar effects of OT in reducing right amygdala responses to all emotional faces, although desensitization effects were only significant for threatening (fear and anger) and happy faces. While some previous studies have found effects of OT only for fear faces (Spengler et al., 2017) or for fear and angry ones (Kirsch et al., 2005), our current findings are in agreement with a number of others reporting that it can reduce amygdala responses to both positive and negative (Domes et al., 2007) or all face emotions (Koch et al., 2016; Quintana et al., 2016). A possible explanation for these differences could be that studies, including our own, using presentation strategies where subjects are not directly required to identify or discriminate between face emotions (Domes et al., 2007; Koch et al., 2016; Quintana et al., 2016) have tended to report valence-independent results. Importantly however, only the magnitude of amygdala responses to fear faces was positively associated with arousal and intensity ratings in the PLC group and OT exerted an anxiolytic action by both reducing these ratings and abolishing their correlation with amygdala activation. Dysregulations in amygdala and behavioral responses to threatening stimuli have been observed across major psychiatric disorders, particularly those characterized by marked social impairments and anxiety (Hennessey et al., 2018; Neumann and Slattery, 2016). There was some evidence that behavioral anxiolytic effects of OT were greater after repeated compared to single doses in the group receiving OT every other day for angry faces but not in terms of amygdala responses. Possibly the short 5 day period for OT-treatment was not sufficient to demonstrate robust enhancement effects of repeated doses and time-dependent enhancement effects of longer duration treatment need to be investigated in studies on clinical populations.

A major finding from our secondary outcome measure is that OT’s attenuations of neural and behavioral responses to face emotions are highly dependent on OXTR genotype. Only G non-carriers of rs53576 and A-carriers of rs2254298 showed significant reductions in amygdala responses to face emotions under OT although only A-carriers of rs2254298 showed evidence for significant reductions in arousal and intensity ratings. A-carriers of both SNPs have frequently been associated with social dysfunction in autism and social anxiety disorders (Cataldo et al., 2018; Jurek and Neumann, 2018). A recent haplotype-based analysis of OXTR SNPs including rs53576 and rs2254298 also indicated an association with sensitivity to OT-effects on face recognition (Chen et al., 2015).

Interestingly, task-related and intrinsic network changes produced by OT showed a strikingly different sensitivity to repeated doses and OXTR genotype and there were no associations between resting state connectivity and either amygdala or behavioral responses to face emotions. However, for the resting state effects of OT there was no advantage of giving OT daily as opposed to every other day. Intrinsic networks may therefore be less sensitive to the effects of repeated OT administration or OXTR genotype than those engaged by external social emotional stimuli, although arguably the effects of OT on neural processing during actual processing of social cues should be of greatest therapeutic relevance.

There are several limitations in the current study. Firstly, we only included male subjects to increase relevance to potential therapeutic use in autism and to avoid menstrual cycle effects. There is however increasing evidence for sex-dependent effects of OT, notably for amygdala responses (Gao et al., 2016; Jurek and Neumann, 2018; Lieberz et al., 2019; Luo et al., 2017) and also for rs53576 (Feng et al., 2015). Thus, females may show different responses to repeated doses of OT and associations with receptor genotype. Secondly, for the genotype analysis, subject numbers are relatively low for establishing robust associations. While treatment x genotype interactions only passed Bonferroni correction for arousal ratings, post-hoc tests for both behavioral and amygdala effects were highly significant.. The AA allele of rs53576 also occurs more frequently in Asian and Caucasian populations (Butovskaya et al., 2016). Finally, we only examined the effects of one dose and time course for the effects of OT and there may be dose-dependent and temporal variations. The 24IU dose and 45 min post-treatment time course used are the same as in many previous studies demonstrating both task and resting-state effects of OT (Kendrick et al., 2017) and maximal effects on reduced amygdala responses to fear faces occur with a 24IU dose from 45-75 min (Spengler et al., 2017). However, higher doses may produce different effects (Spengler et al., 2017) and a recent regional cerebral blood flow study has reported reduced resting amygdala activity at shorter time points following 40IU intranasal OT (Martins et al., 2020).

In conclusion, the current study provides the first evidence for an important influence of OT dose frequency and receptor genotype on its modulation of neural and behavioral responses to emotional faces in healthy human subjects. Dose frequency therefore requires further empirical evaluation in patient populations given that it can critically determine treatment efficacy in clinical trials employing chronic administration.

## Data Availability

All data referred to in the manuscript will be available

## Acknowledgement

We thank the reviewers and editor for their comments and suggestions for improving the manuscript.

## Author contributions

JK and KMK designed the experiment. JK, YZ and CS carried out the experiment. JK, KMK, CM, FZ and BB analyzed the experiment and JK, KMK, CM and BB wrote the paper. All authors contributed to the conception of the study and approved the paper.

